# Baptism of fire: Modeling the effects of prescribed fire on tick-borne disease

**DOI:** 10.1101/2022.01.01.21268589

**Authors:** Emily Guo, Folashade B. Agusto

## Abstract

Recently, tick-borne illnesses have been trending upward and are an increasing source of risk to people’s health in the United States. This is due to range expansion in tick habitats as a result of climate change. Thus, it is imperative to find a practical and cost-efficient way of managing tick populations. Prescribed burns are a common form of land management that can be cost efficient if properly managed and can be applied across large amounts of land. In this study, we present a compartmental model for ticks carrying Lyme disease and uniquely incorporate the effects of prescribed fire using an impulsive system to investigate the effects of prescribed fire intensity (high and low) and the duration between burns. Our study found that fire intensity has a larger impact in reducing tick population than the frequency between burns. Furthermore, burning at high intensity is preferable to burning at low intensity whenever possible, although high intensity burns may be unrealistic due to environmental factors. Annual burns resulted in the most significant reduction of infectious nymphs, which are the primary carriers of Lyme disease.

## 1 Introduction

Many ticks are disease vectors that significantly impact public health. Reports of overall tick-borne diseases doubled from 2006 to 2018 [3] while the incidence of Lyme disease in the United States has been steadily increasing, from a little less than four cases per 100,000 people in the 1990s to close to 10 cases per 100,000 people in the early 2000s [21]. New pathogens continue to emerge, including heartland virus, *Bourbon virus, Borrelia miyamotoi, Borrelia mayonii*, and *Ehrlichia muris eauclairensis* [10]. Climate change has expanded the northern borders of tick habitats and increased winter tick activity, contributing to the prevalence of tick-borne diseases [11]. Therefore, finding a practical and cost-efficient way to manage tick populations has become extremely important. The majority of ticks that carry Lyme disease are infected through mice or other small rodents [2], so most methods that have looked at tick reduction are focused on either host reduction or tick elimination [20]. Ticks can have up to 2 to 3-year long life cycles, with four general stages that each require one blood meal after the eggs hatch: egg, larvae, nymph, and adult [20]. The majority of human infections come from tick nymphs, which are much smaller than adult ticks (less than 2mm long or about the size of a poppyseed), making them more difficult to spot on the human body and therefore more likely to remain undetected [20]. Nymphs are also more numerous than adult ticks and are most active during the spring and summer months, when the number of people who spend time outside is substantially larger than those during other months. The blacklegged tick, also known as Ixodes scapularis, has a life-cycle that generally lasts two years, while the life-cycle of the lone star tick (*Amblyomma americanum*) is around three years long. Most ticks hatch from their eggs in the spring and have the ability to live for three to five months between each blood meal [9].

Prescribed fires, or controlled burns, are a common and necessary form of land management in many different environments that are also effective in controlling tick populations. This is through both directly killing ticks along with destroying their leaf-litter habitat [9]. Larvae, nymphs, and adults spend the vast majority of their time in leaf litter other than the few days that they are feeding on their hosts. Controlled burns are appealing due to their time and cost efficiency along with their ability to be applied across a large amount of land. They are generally most effective in the late spring and early summer, as that time coincides with when nymph ticks are questing for hosts (although this is heavily dependent on the type of land that is being burned) [5]. Primary concerns around prescribed fire include air quality (due to smoke) and the potential for the fire to burn out of control; however, these can be prevented when proper precautions are taken.

Many studies have looked at the impact of prescribed fires on tick populations, with conflicting results. The majority of these studies agree that tick populations decrease immediately after a burn but recover to pre-burn abundance after around one year [20]. Other studies have found that although the nymph population decreased, the risk of encountering infectious nymphs remained the same [16] or that the tick population even increased [19]. However, these studies often fail to account for the logistics of true prescribed burning (long term and over lots of land on a regular basis) or other predictors of tick abundance such as host abundance, climate, or vegetation structure [20]. There is also the possibility that post-burn recolonization rates vary based on tick species, habitat type, climate, and burn intensity [1, 6].

A study done by Allan [1] in the oak-hickory ecosystems of the Missouri Ozarks looked at the relationship between lone star tick larvae populations and deer abundance under long-term burn management. The sites were burned in the spring at low intensity every 3-5 years. The ticks were depleted but then rapidly grew starting two years post-burn, coming back down to pre-burn abundance around five years post-burn [1]. The researchers attributed this increase to the high host populations post-burn, as freshly burnt areas are better for deer to forage in. These issues could be countered by more frequent, longer, and larger scale burns, which correlates with other studies that also believe that burns at higher intensity are most effective in countering ticks than those at low intensity [8]. Gleim *et al* [10] found that long term prescribed fire (regular burning for 10+ years) significantly reduced tick abundance, regardless of burn interval, host abundance, or vegetation structure. This is primarily due to the change in vegetation structure, creating a hotter and drier environment that is less appealing for ticks [20]. These burnings decreased the encounter rate with infectious ticks by 98% in plots in southwestern Georgia and northwestern Florida. However, more research in a variety of environments needs to be done regarding realistic prescribed burning as a tick management technique.

The goal of this study is to develop a compartmental model for ticks carrying Lyme disease to see how they are affected by prescribed burns. We look at both fire intensities (high and low) and the duration between fires in order to understand how this common land practice affects tick populations and the prevalence of Lyme disease among them. We also investigate whether intensity or duration plays a more significant role in tick population reduction overall. To the best of our knowledge, this is the first study to use a mathematical model for Lyme disease to examine the effects of prescribed fire.

The remainder of the work in this paper is organized as follows. In Section 2, we formulate our baseline tick/Lyme disease model, compute the model basic reproduction number, and carry out basic stability analysis including sensitivity analysis to determine the parameter with the most impact on the basic reproduction number. In Section 2.3, we describe the tick model with the effect of prescribed fire using an impulsive system of ordinary differential equations and present some stability analysis results of the impulsive system. In Section 2.3.1, we discuss the estimation of parameters related prescribed fire from literature. In Section 3, we present some simulation results, and in Section 4 we discuss our findings and close with conclusions.

## 2 Materials and Methods

This model was created by incorporating two subgroups: mice and ticks. The mice population is divided into susceptible (*S*_*M*_ (*t*)) and infected mice *I*_*M*_ (*t*)). The tick population is divided by life stage (eggs, larvae, nymph, and adult) and further divided into susceptible and infected groups for larvae (*S*_*L*_(*t*) and *I*_*L*_(*t*)), nymphs (*S*_*N*_ (*t*) and *I*_*N*_ (*t*)), and adults (*S*_*A*_(*t*) and *I*_*A*_(*t*)). Since ticks must take a blood meal before they become infected and there is no vertical transmission for the disease, all eggs remain susceptible (*S*_*E*_(*t*)). Individuals move between compartments according to their life stage and disease status. We assume that all transition rates are of the current population and remain steady, with no migration into or out of the overall population.

The force of infection in the mice (or the rate that susceptible mice become infected) is given as

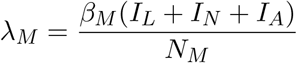

where the parameter *β*_*M*_ is the probability that infection will occur if a mouse is bitten by an infectious tick, multiplied by the number of all infectious ticks – the sum of the larvae, nymphs, and adults – and then divided by the total number of mice where *N*_*M*_ = *S*_*M*_ + *I*_*M*_. For simplicity, we assume that there is a homogenous mixing of both mice and tick populations. New mice are born at a rate of *π*_*M*_, and the susceptible mice move into the infected compartment at a rate of *λ*_*M*_. Since the disease does not affect the mice, they remain in the infected compartment for the rest of their lives and death related to the disease is not incorporated into the model. Therefore, natural death, given as the rate *μ*_*M*_, is the only factor decreasing the population of both susceptible and infected mice. The equations for the susceptible mouse population and infected mouse population are shown below:

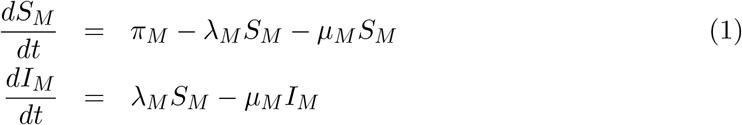

The force of infection in the ticks (or the rate that ticks become infectious) is given as

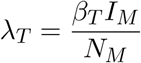

where the parameter *β*_*T*_ is the probability that infection will occur if a tick bites an infected mouse, multiplied by the total number of infected mice and divided by the overall mice population (both susceptible and infected). Since there is no vertical transmission of the disease between adult ticks and their eggs, we assume that there is no infected egg compartment. We also assume that all adult ticks are capable of reproduction, regardless of whether they are susceptible or infected. The eggs mature into the larvae category at a rate of *σ*_*T*_, with a certain percentage dying naturally at the rate *μ*_*E*_.

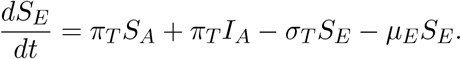

The susceptible larvae then have the possibility of moving into the infected larvae category at a rate of *λ*_*T*_ or remaining in the susceptible larvae compartment. Both susceptible and infected larvae populations are affected by the natural death rate of *μ*_*L*_. They also both move into their respective nymph compartments at a rate of *τ*_*T*_, regardless of whether they are susceptible or infected. This leads to the following system of equations for larvae

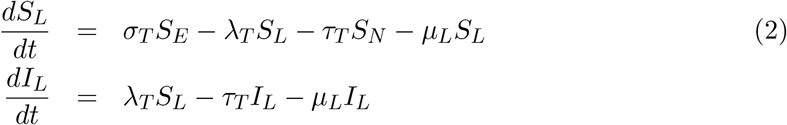

After taking another blood meal, susceptible nymphs move into the infected nymph compartment at the rate *λ*_*T*_. Both susceptible and infectious nymph populations are reduced by the natural death rate of *μ*_*N*_ and continue to mature into adults at the rate of *γ*_*T*_. The equations for the development rate of nymphs are given below

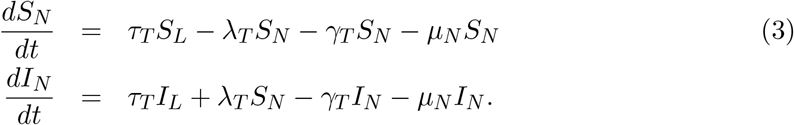

Although it is less common than at other life stages, adult ticks are still able to become infectious. Susceptible adults move into the infected compartment at the rate *λ*_*T*_, after they take a blood meal at that life stage. Both susceptible and infected adult populations are removed by natural death rate of *μ*_*A*_. The equations for both susceptible and infected adult ticks are given as

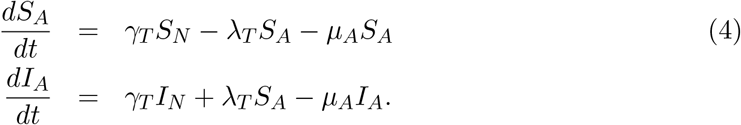

Incorporating all the assumptions and equations above, we have the following system of differential equations:

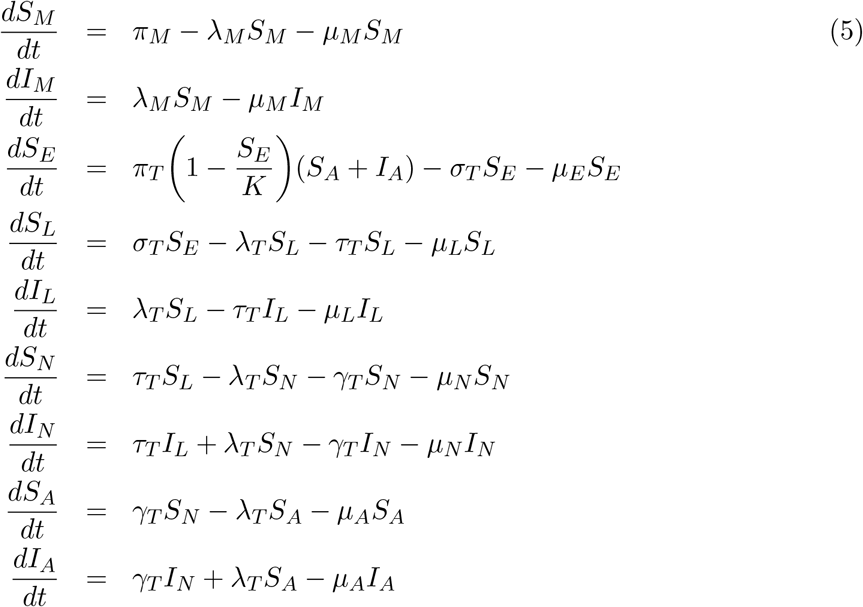

The related model schematic is given in Figure 1 and the description of the model variables and parameters are stated in Table 1.

**Table 1:** Description of the variables and parameters for the Lyme disease model (5).

| Variable | Description |
| --- | --- |
| $S_M(t)$ | Number of susceptible mice |
| $I_M(t)$ | Number of infected mice |
| $S_E(t)$ | Number of eggs |
| $S_L(t)$ | Number of susceptible larvae |
| $I_L(t)$ | Number of infected larvae |
| $S_N(t)$ | Number of susceptible nymphs |
| $I_N(t)$ | Number of infected nymphs |
| $S_A(t)$ | Number of susceptible adults |
| $I_A(t)$ | Number of infected adults |

| Parameter | Description |  |  |
| --- | --- | --- | --- |
| $\pi_M$ | Mice birth rate | 0.02 | [15] |
| $\mu_M$ | Mice mortality rate | 0.01 | [15] |
| $\beta_M$ | Tick-to-mouse transmission probability | 0.9 | [13] |
| $\pi_T$ | Tick birth rate | 456.36 | [15] |
| $K$ | Carrying capacity | 5,000 | [14] |
| $\mu_E$ | Death rate / inviability rate of the egg | 0.0025 | [15] |
| $\sigma_T$ | Eggs to larvae developmental rate | 0.00677 | [15] |
| $\mu_E, \mu_L, \mu_N, \mu_A$ | Tick mortality rate at different life-stages | | |
| $\mu_T$ | Tick mortality rate | 0.015 | [15] |
| $\beta_T$ | Mouse-to-tick transmission probability | 0.9 | [13] |
| $\tau_T$ | Larvae to nymphs development rate (or nymph development rate) | 0.00618 | [15] |
| $\gamma_T$ | Nymphs to adults development rate of (adult development rate) | 0.00491 | [15] |

**Figure 1:**
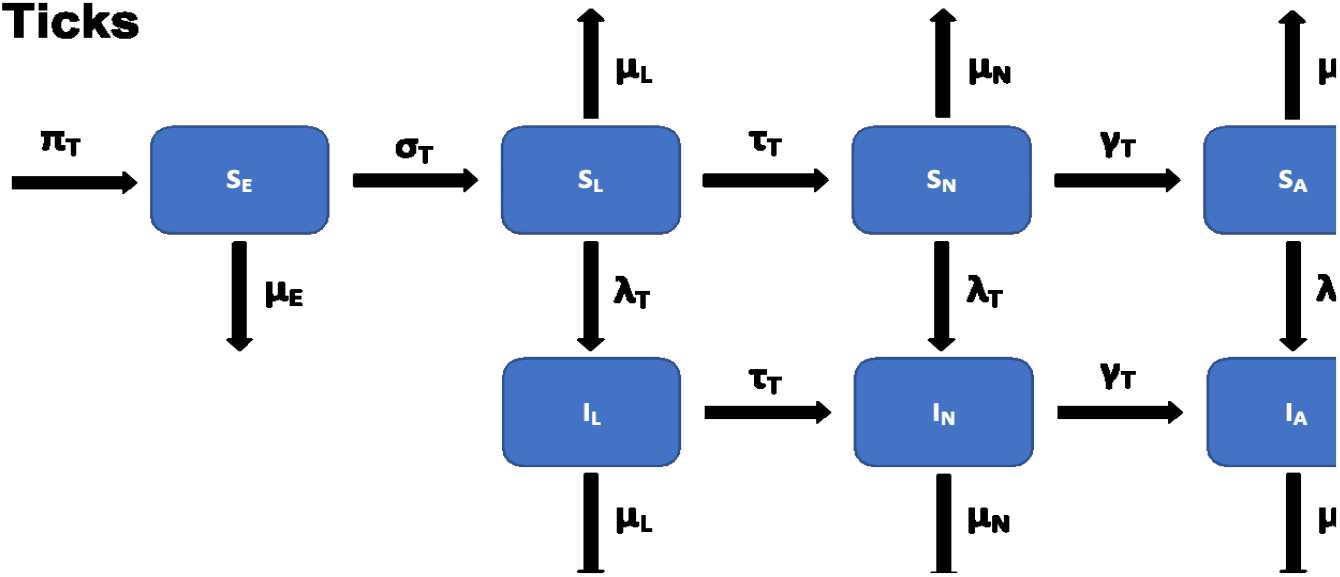
Flow diagram of the Lyme disease model for ticks and mice. The mice population is represented by the red boxes and is divided into susceptible (*S*_*M*_ (*t*)) and infected (*I*_*M*_ (*t*)) compartments. The tick population is represented by the blue boxes and is divided by life stage and infection status. It consists of susceptible eggs (*S*_*E*_(*t*)), susceptible larvae (*S*_*L*_(*t*)), infected larvae (*I*_*L*_(*t*)), susceptible nymphs (*S*_*N*_ (*t*)), infected nymphs (*I*_*N*_ (*t*)), susceptible adults (*S*_*A*_(*t*)), and infected adults (*I*_*A*_(*t*)).

The basic qualitative properties of the tick model (5), its positivity, and the boundedness of solutions are given in Appendix A

### 2.1 Reproduction number *ℛ*_0_

The associated reproduction number using the next generation matrix method[4, 17] for the Lyme disease model (5), denoted by *ℛ*_0_, is given by

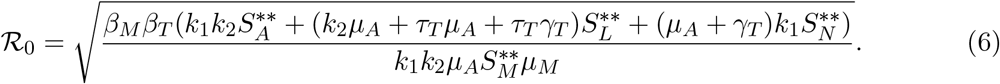

However we made a simplifing assumption that *μ*_*L*_ = *μ*_*N*_ = *μ*_*A*_ = *μ*_*T*_. Then, the reproduction number in (6) becomes

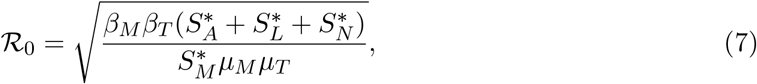

see details in Appendix B.

The basic reproduction number, *ℛ*_0_, is defined as the expected number of new infections that result from one infectious individual in a population that is fully susceptible [4, 17]. This value is extremely significant because if the reproduction number is less than unity (*ℛ*_0_ *≤* 1) then the disease cannot invade the population and it will die out in the community. Conversely, if *ℛ*_0_ > 1 then the disease will continue to persist in the population. This determines whether there is a possibility of disease elimination or if the goal should be to manage transmission within the community.

### 2.2 Sensitivity analysis

Sensitivity analysis was conducted in order to determine the contribution of each of the model parameters on the reproduction number *ℛ*_0_. Results of this help identify which parameters are the best to target regarding interventions and future data collection. A normalized forward sensitivity index was used to determine the ratio of the relative change in *ℛ*_0_ based on a relative change in a parameter. The sensitivity indices of *ℛ*_0_ is derived as

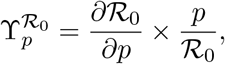

where 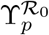 is the forward sensitivity index of *ℛ*_0_ with respect to parameter *p*. Parameter *p* is a parameter within *ℛ*_0_. The results of this sensitivity analysis are shown below in Figure 2:

**Figure 2:**
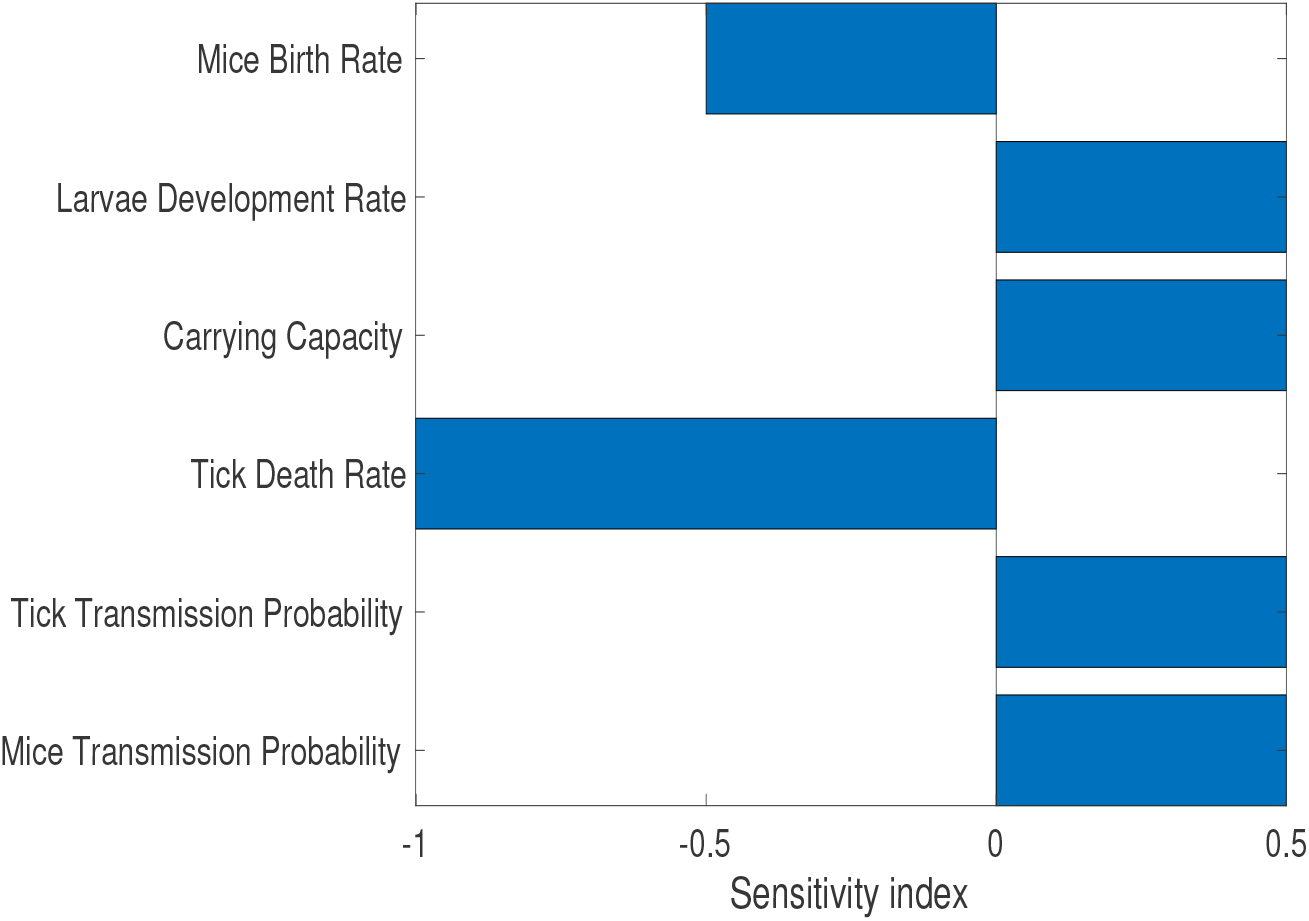
Sensitivity analysis results for the Lyme disease model. The larvae development rate, the carrying capacity, and the disease transmission probability for both mice and ticks are all positive influences on *ℛ*_0_. The mice birth rate and tick death rate are negative influences on *ℛ*_0_. Prescribed fire directly increases the tick death rate, making it an effective control strategy.

The larvae development rate, the carrying capacity of the environment, and the disease transmission probability for both mice and ticks all positively affect *ℛ*_0_. As these values increase, *ℛ*_0_ will also increase. We are more interested in the mice birth rate and the tick death rate, which negatively affect *ℛ*_0_. As these parameters decrease, *ℛ*_0_ will also decrease. The most influential parameter on *ℛ*_0_ is the tick death rate. This suggests that control strategies that effectively target the spread of Lyme disease will focus on increasing the tick death rate, and to a lesser extent, also decreasing the mice birth rate. The sensitivity analysis aligns with our results from the model, as prescribed fire is a mechanism that most significantly affects the tick death rate.

### 2.3 Tick model with prescribed fire

In this section we consider the effect of fire on ticks and the small mammal population. We do not explicitly incoporate fire into the model (5); rather, we consider the effect of fire on population size after the burns. To introduce the effect of prescribed fire into the Lyme disease model (5), we have the following system of non-autonomous impulsive differential equations.

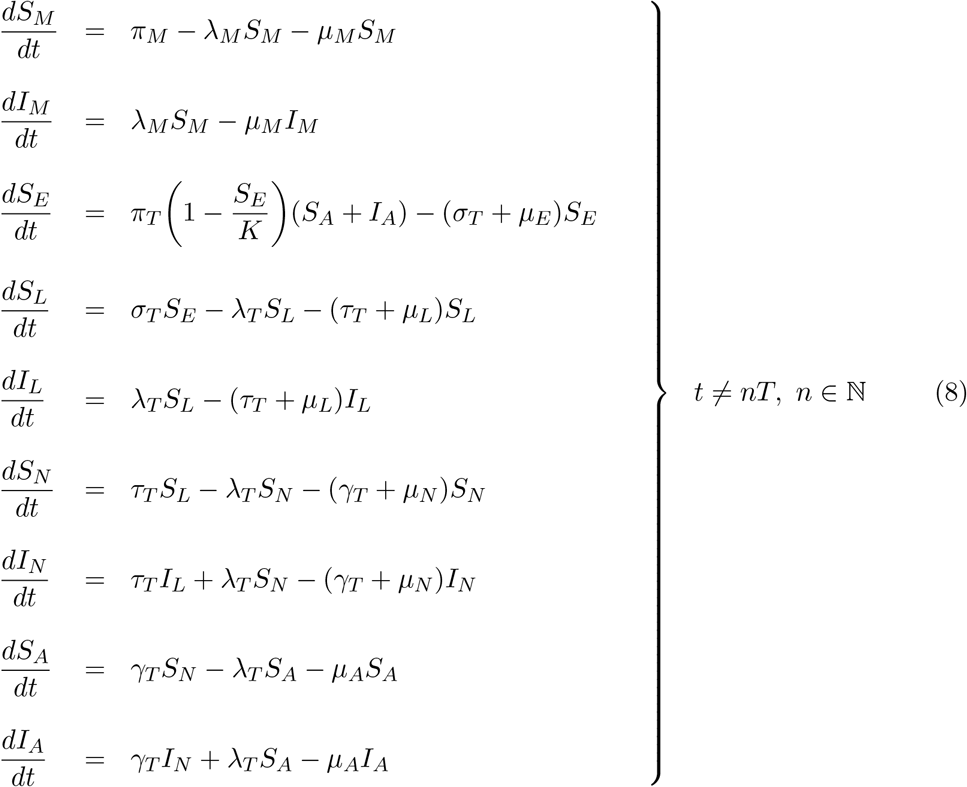

subject to the prescribed fire impulsive condition

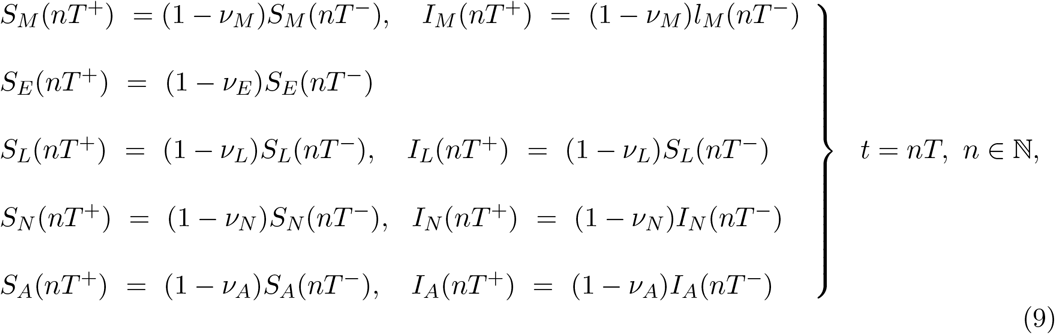

where *t*_*n*_ is the times that prescribed fire is implemennted, which may be fixed or non-fixed; in this study we will consider the case with fixed times. The parameters *ν*_*j*_, where *j* = *E, L, N, A, M* are the proportion of the tick and mice population that is reduced by the fire. In Section 2.3.1 below, we discuss how these parameters are estimated using data from low and high intensity fires. The existence, and stability of the impulsive model (8) are given in Appendix C.

#### 2.3.1 Prescribed fire parameter estimation

To estimate the parameters (*ν*_*L*_, *ν*_*N*_, *ν*_*A*_, *ν*_*M*_) which quantify the reduction in each tick life-stage and mice population after the different burn intensities (low and high burns) we use data from [8, 18]. The parameters for each life stage relating to the low intensity burn were estimated using data from [8], while the parameters relating to the high intensity burn were estimated using data from [18]. Each group of parameters was separated into the larvae, nymph, and adult life stages, but only data in [18] provided data to estimate the parameter for mice population. Below we give a summary description of the study sites in each study, the amount of ticks and mice collected and how these parameters are estimated from the data collected.

##### High intensity fire

The study in [18] was conducted in chaparral habitat at the University of California, Hopland Research and Extension Center, in Mendocino County, CA. The study took advantage of two prescribed fires ignited on 1 June 1995 that were intended to reduce fire load in two chaparral plots, Maude’s Glade (MG) and Don’s Brush Plot (DBP). The fires were ignited by hand crews using drip torches in a strip headfire configuration to produce relatively uniform fire behavior that left no live branches in the shrub-line, which we assume as high intensity fire. For a period of 13 months beginning a month before the burn, control and treatment areas were monitored for the presence of ticks by flagging the vegetation or ground between 0800 and 1000 hours, along with CO_2_-baited pitfall traps. In order to assess the abundance of rodents and the associated ticks on them, live traps were set to catch the rodents.

At the two study sites (MG and DBP), six tick species (namely, *Ixodes pacificus, Ixodes jellisoni, Ixodes spinipalpis, Ixodes woodi, Dermacentor occidentalis* and *Dermacentor parumapertus*) were removed from the six different rodents species caught; these include California kangaroo rat (*Dipodomys californicus californicus*), brush mouse (*Peromyscus boylii*), pin*o*∼n mouse (*P. truei sequoiensis*), deer mouse (*P. maniculatus gambelii*), dusky-footed woodrat (*Neotoma fuscipes*), and western harvest mouse (*Reithrodontomys megalotis longicaudus*). After the fire treatment, about half as many rodents were trapped at the treated sites compared with control sites.

All the ixodid tick species (*Ixodes pacificus, Ixodes jellisoni, Ixodes spinipalpis, Ixodes woodi*) are competent hosts that are able to transmit Lyme disease, but we use only the data of *Ixodes pacificus* to estimate the parameter used to quantify the reduction in the tick population as a result of fire. Table 2 gives a break down of *Ixodes pacificus* collected on the rodents in the control and treated areas at both MG and DBP sites, as well as the number rodents collected at these sites pre-and post-burn.

**Table 2:**
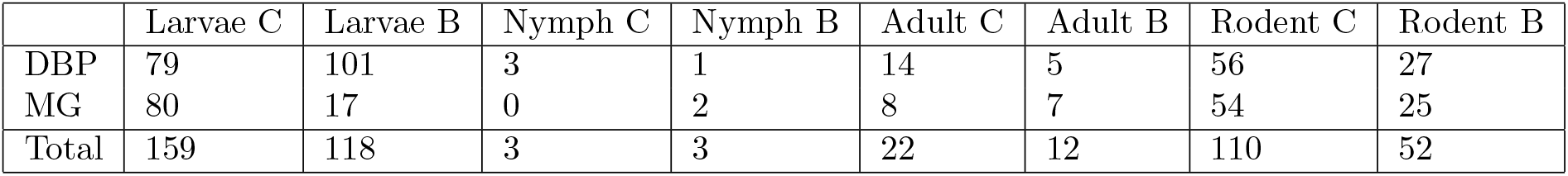
Data taken from [18] under assumed high intensity fire. C = control, B = burn.

##### Computing parameters *ν*_*L*_, *ν*_*N*_, *ν*_*A*_, *ν*_*M*_ for high intensity fire

To compute these parameters we assume that equal numbers of ticks and rodents are in the sites (control and treated sites) pre-burn, and the difference in number is due to the burn, since we have no way to measure exactly the number of ticks in all the study sites. First, we take the difference between the total number of ticks and mice in the control and treatment sites and divide it by the total number of ticks and mice in the control sites. Then we subtract these proportions from 1 to give the proportions reduced as a result of the burn.

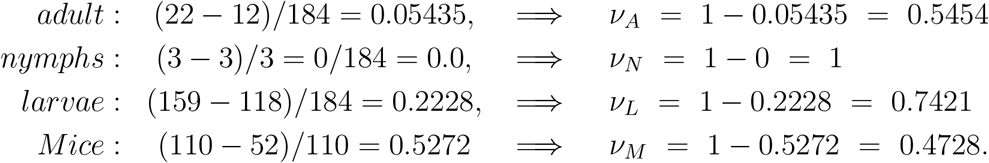

##### Low intensity fire

The study in [8] was conducted in an open oak woodland barren complex atWestern Illinois University’s Alice L. Kibbe Field Station located in Warsaw, in Hancock County, IL, USA. These areas are comprised of multiple habitat types including oak-hickory woodlands, early successional woodlands, oak barrens, floodplain forests, restored tallgrass prairies and hill prairies. The entire study site was last burned in 2004 (B04) and two additional burns were carried out in spring of 2014 (B14) and 2015 (B15). The burns were considered low intensity because most flame heights were less than 1 meter and plant mortality was limited to the understory vegetative community [23].

Ticks were collectted through flagging method every two weeks when the vegetation was dry between 1200 and 1800 hours, during two consecutive years (9 May 2015 until 30 October 2015 and 22 April 2016 until 4 November 2016). A total of 2788 *Amblyomma americanum*, 54 *Ixodes scapularis*, and 23 *Dermacentor variabilis* ticks were collected in 2015 and 2016. *Amblyoma americanum* ticks collected in B04 made up 51% of the collection (*n* = 1433), while those collected in B14 made up 37% (*n* = 1045) of the collections, and those collected in B15 constituted 11% (*n* = 307) of the collection. Of these ticks, 2% (*n* = 67) were adults, 4% (*n* = 107) were nymphs, and 93% (*n* = 2614) were larvae. Of the 23 *D. variabilis* collected, 74% (*n* = 17) were adults, 9% (*n* = 2) were nymphs, and 17% (*n* = 4) were larvae. While 4% (*n* = 2) of the 54 *I. scapularis* collected, were adults, 22% (*n* = 12) were nymphs, and 74% (n = 40) were larvae. Here we use the data collected for *Ixodes scapularis* to estimate the parameter quantifing the reduction in the tick population due to fire since *I. scapularis* can transmit Lyme disease. The study did not indicate if these were data for the pre-burn or post burn number of *I. scapularis* collected, nor did it provide data for the number of mice that were caught.

##### Estimating parameters *ν*_*L*_, *ν*_*N*_, *ν*_*A*_, *ν*_*M*_ for low intensity fire

To estimate these parameters, we assume the 54 *I. scapularis* collected are the ticks left after the burn. We then divide the numbers collected in each age group by the total number collected and subtract the proportion obtained from 1 to obtain the proportion reduced by fire.

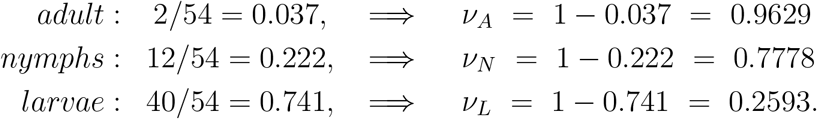

Although, these were the best sources of data we could find (that accounted for tick life stages pre- and post-burn along with fire intensity); unfortunately, they were not ideal. The burns were conducted in vastly different environments – the high intensity fire took place in California chaparral while the low intensity fire took place in Illinois oak woodland. The high intensity burn was only conducted once in the summer, while the low intensity burn was conducted in the spring for two consecutive years. We were unable to find any sources of data from similar geographic locations or number of burns that were detailed enough in terms of tick data and fire intensity data to be used. These variances between the two data sets have the potential to drastically affect the burn results.

## 3 Results

To address our research goals, we started by looking at how different burn frequencies and intensities affected tick populations, focusing specifically on the nymphs; since the primary mode of transmission for Lyme disease from ticks to humans is through infectious nymphs. High intensity fires substantially change the above ground structure, with no live branches and few shrub skeletons left over [6, 23]. These burns are much more uniform than those at lower intensities, which are patchy and have vegetation cover and woody debris left over that ticks are able to survive the burn in [8]. Our first simulation in Figure 3 shows the substantial difference between high intensity and low intensity burns regarding how effective they are at reducing the infectious nymph population. Although both burns start out at around the same effectiveness regardless of intensity, the high intensity burn proves to reduce the infectious nymph population much more efficiently than the low intensity burn. The infectious nymph population affected by the low intensity fire continues to increase despite the burns, while the infectious nymph population affected by the high intensity fire remains consistently low.

**Figure 3:**
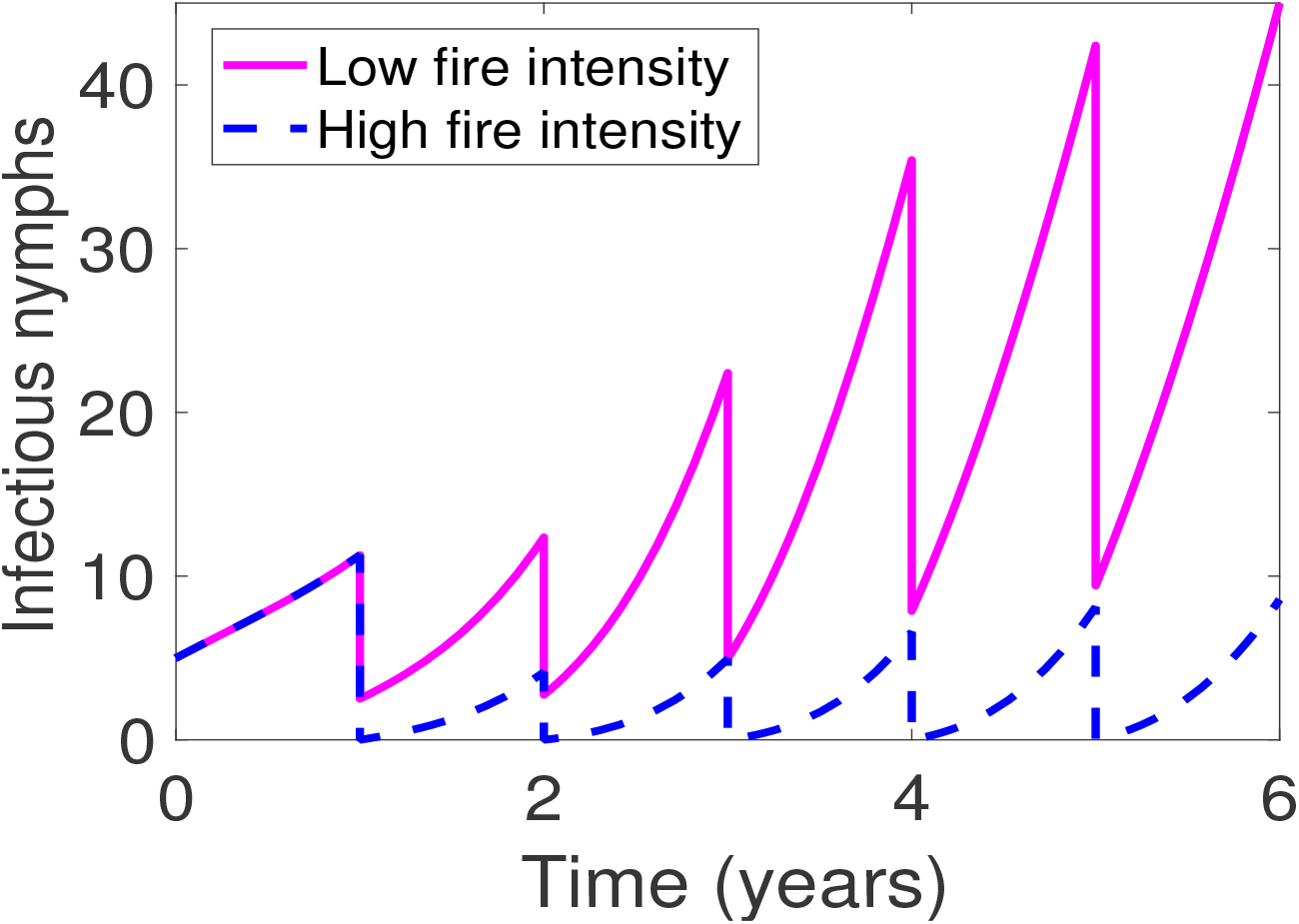
Simulation of annual burns for six years at varying intensities. Parameter values are given in Table 1.

In our second simulation shown in Figure 4, we consider the duration between burns for both high and low intensity fires. We consider the effects of burning for a period of six years once every six years, once every three years, once every other year, and annually. We found in general that as the duration between burns decreases the infectious nymph population also decreases regardless of the burn intensities. Furthermore, we found that the duration between burns has a more significant effect on ticks with higher intensity fires than with lower intensity fires. Fire intensity appears to have a larger influence on tick reduction than duration of the burns, as burning fewer times at a higher intensity is more effective than burning more times at a lower intensity. For example, high intensity burns once every three years reduces the infectious nymph population more than low intensity burns once every two years. However, high intensity burns might be unrealistic due to environmental factors. In that case, annual burns at low intensity result in the most significant reduction of infectious nymphs in its category.

**Figure 4:**
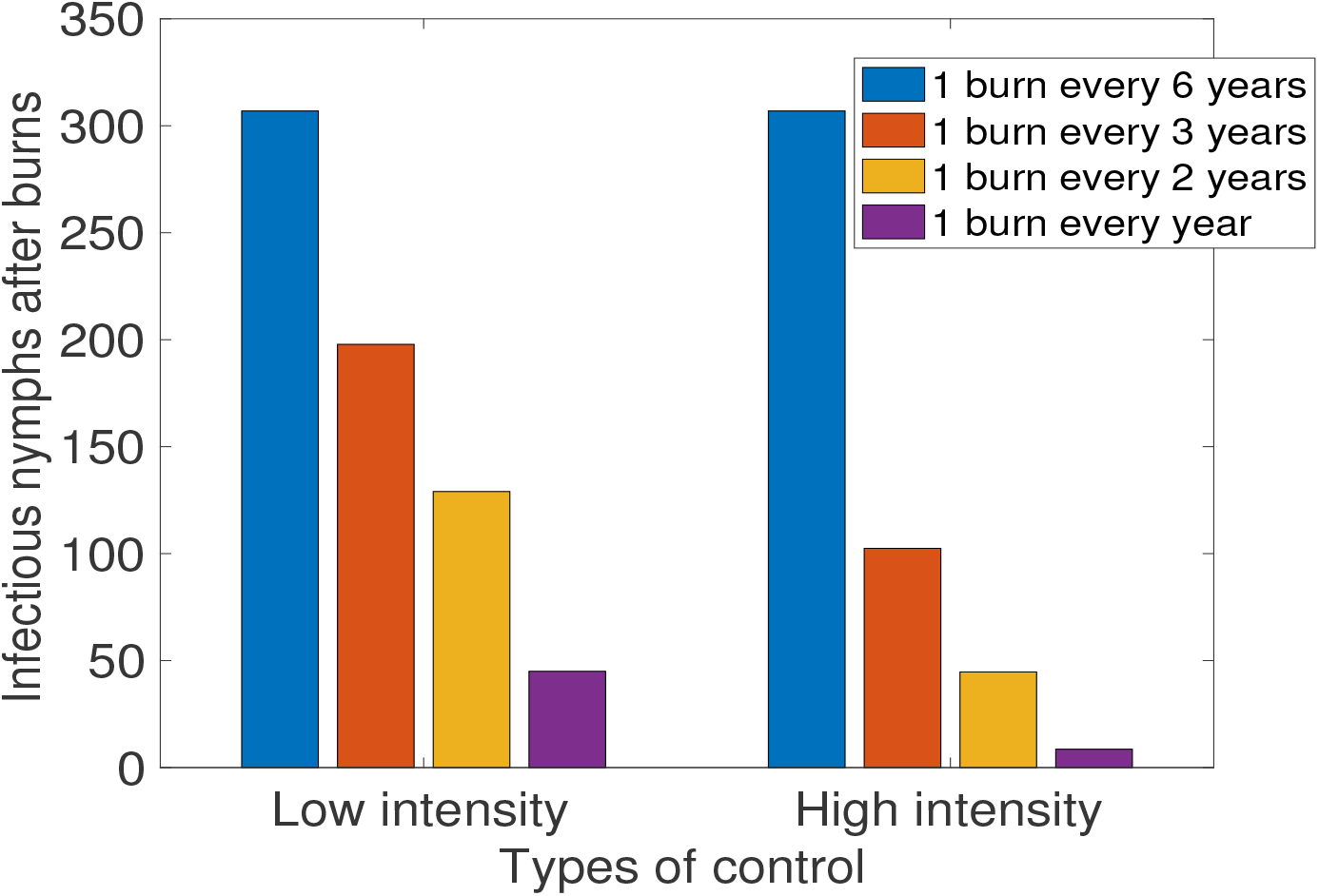
Simulation of varying intervals between burns, separated by their intensity

## 4 Discussions and conclusions

### 4.1 Discussions

The most effective simulation in minimizing infectious nymph populations is having annual high intensity burns, although this is not always practical due to environmental factors such as weather, burn location, and fuel loads. The geographic location of the burn is also important to consider, as it is riskier to have higher intensity fires closer to human settlements. Most prescribed fires are at low to moderate intensity and are repeated every 1-5 years, with their primary objective being to reduce fuel loads and act as a wildfire prevention tool [18]. There have been a few negative outcomes associated with intense annual burning such as oak tree mortality, soil compaction, and an increased number of tree cankers and tree colonization by root fungus [12], but these depend heavily on the type of environment that is being burned. Overall, infectious tick populations substantially decrease as the time between burn also decreases, regardless of the intensity that the burn is conducted at.

These findings were confirmed in a private conversation with Gallagher of the U.S. Forest Service Research and Development team. He recently finished working on a study that looked at how long-term prescribed burns affected tick populations in New Jersey oak woodlands. They found preliminary data that strongly suggests that different tick species are impacted differently by prescribed burns due to their varying moisture sensitivities [6]. It appears that black-legged ticks, the primary vector of Lyme disease in humans, are the most sensitive to moisture loss while Lone Star ticks and Gulf coast ticks seem to be more resilient. Gallagher believes that one of the reasons for this is the thickness of the scutum, which is a shield-like plate on the back of hardback ticks [6]. In general, initial data from this study agrees with the results of our model. Their high intensity burns, occurring once every 20 years with data collection for four consecutive years after the burn, saw the greatest reduction in tick populations. Annual low intensity burns for over 25 years had the second largest tick reduction, while the single low intensity burn showed the smallest amount of tick reduction.

Few research studies have examined the effects of prescribed fire on tick populations and disease prevalence [1]. The studies that do often choose to focus on one aspect, usually that of how the burns affect the ticks, with the fire itself being of secondary interest [9]. Future research that records detailed information about fires, such as flame height, difference in vegetation presence before and after the burn, how the fire was ignited, weather conditions during the burns, overall fire behavior, etc. would be extremely helpful in ensuring the accuracy of the parameters used in the model. Knowledge about the exact dates of the burns and the specifics of the duration between burns is also useful, as seasonality has the potential to play a large role in the effectiveness of burns on disease prevalence. This is especially true with diseases that are primarily found to be transmitted to humans in a certain life stage of the tick, such as with nymphs and Lyme disease. Knowing the exact dates of the burns along with the time in between is useful when comparing the burn results to tick life cycles. For example, spring burns will mostly kill nymphs, although this can vary by species. Noting the geographic context is also important, as environment plays a role in how these burn impact tick populations. Finally, increasing the time frame of studies that look at how prescribed fire impacts ticks will also be useful. Many studies that claim to study how ticks are affected by prescribed fires only involve a single burn, which is vastly different from true prescribed burning, which involves regular fires over many years.

Land geography seems to have a role in the effectiveness of prescribed burns on tick population reduction regarding how fast the environment is able to regrow back to its original state before the burn, although to the best of our knowledge there have not been any studies that specifically look at this. Certain environmental factors might make certain geographic locations better candidates for prescribed fires when the main goal is to reduce tick populations, and this is a subject that is worth looking into in the future. It also appears that different tick species have different levels of heat and moisture resistance, causing them to be affected differently by the fires [6, 8]. Only very preliminary research has been done on this so far, although it is a topic that will substantially influence the choice of whether or not to use prescribed fires for tick reduction.

### 4.2 Conclusion

To conclude, in this study we develop a simple model for Lyme disease transmission and used it to investigate the impact of prescribed burn frequency and fire intensity on the spread of Lyme disease. To the best of our knowledge, this is the first model to incorporate the effect of fire into a mathematical model of ticks and Lyme disease,. The key findings from this study are summarized below.

The simulations of the Lyme disease model (5) with prescribed burns show that:

i. The most influential parameters impacting the reproduction number, *ℛ*_0_, from the sensitivity analysis are *π*_*M*_, *σ*_*T*_, *K, μ*_*T*_, *β*_*T*_, *β*_*M*_.
ii. Intensity appears to be a larger influence on tick reduction than duration is – it is better to burn fewer times at a high intensity than to burn more often at a lower intensity.
iii. Burning at high intensity is preferable to burning at low intensity whenever possible, although high intensity may be unrealistic due to environmental factors.
iv. For low intensity, annual burns resulted in the most significant reduction of infectious nymphs, which are the primary carriers of Lyme disease.

This study has shown the effect of prescribed fire on ticks and the spread of Lyme disease. In a future study we will consider the impact of seasonality on the effectiveness of prescribed burns and the impact of the timing of the burns on disease spread. Geographic landscape and tick habitat are important factors affecting the tick population. In another future work, we will consider different types of geographic landscapes and determine which landscape is most effective in using prescribed burns for tick population control and reduction.

## Data Availability

All data used in this study came from published, cited sources, and are included in the text.

## Data Availability

All data used in this study came from published, cited sources, and are included in the text.

## Acknowledgement

This research was supported by National Science Foundation EPSCOR Track 2 under the grant number 1920946.

## A Analysis of the tick model (5)

### Basic qualitative properties

#### Positivity and boundedness of solutions

For the tick model (5) to be epidemiologically meaningful, it is essential to prove that all its state variables are non-negative for all time. In other words, solutions of the model system (5) with non-negative initial data will remain non-negative for all time *t* > 0.

##### Lemma 1.

*Let the initial data F* (0) *≥* 0, *where F* (*t*) = (*S*_*M*_ (*t*), *I*_*M*_ (*t*), *S*_*E*_(*t*), *S*_*L*_(*t*), *I*_*L*_(*t*), *S*_*N*_ (*t*), *I*_*N*_ *I*_*A*_(*t*)). *Then the solutions F* (*t*) *of the tick model* (5) *are non-negative for all t* > 0.

*Proof*. Let *t*_1_ = sup {*t* > 0 : *F* (*t*) > 0 *∈* [0, *t*]}. Thus, *t*_1_ > 0. It follows from the first equation of the system (5), that

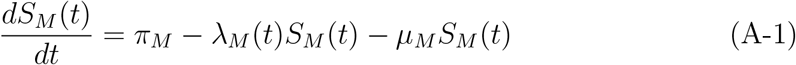

where 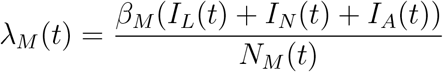

Using the integrating factor method we can rewrite equation (A-1) as

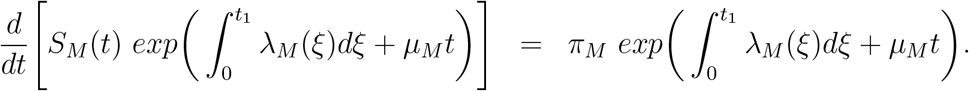

Hence,

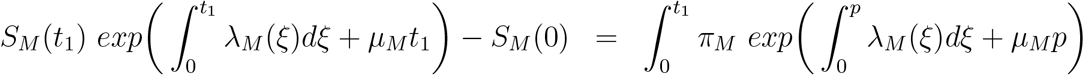

so that,

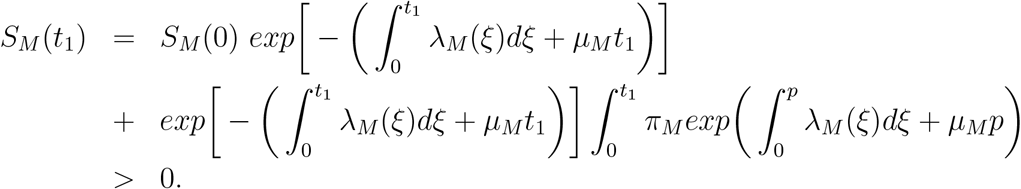

Similarly, it can be shown that *F* > 0 for all *t* > 0.

#### Invariant regions

The Lyme disease model (5) will be analyzed in a biologically-feasible region as follows. Consider the feasible region

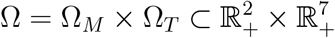

where,

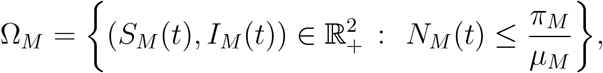

and

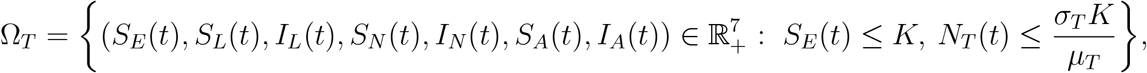

where *N*_*T*_ = *S*_*L*_ + *I*_*L*_ + *S*_*N*_ + *I*_*N*_ + *S*_*A*_ + *I*_*A*_.

##### Lemma 2.

*The region* 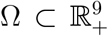 *is positively-invariant for the model (5) with nonnegative initial conditions in* 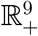.

*Proof*. Summing the first two equations of model (5), we have

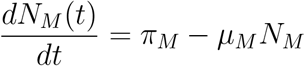

Thus,

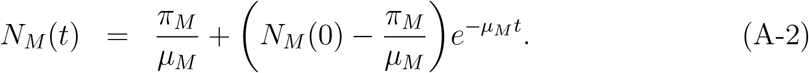

In particular, if 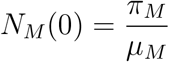, then 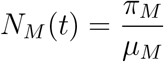.

Next, the last seven equations of model (5) give the following after summing the equations representing the larvae, nymphs, and adult stages

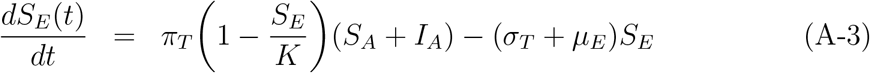

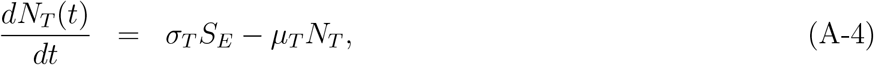

where *μ*_*T*_ = min {*μ*_*L*_, *μ*_*N*_, *μ*_*A*_}. Since *K* is the carrying capacity, it follows that *S*_*E*_ *≤ K*. Hence, equation (A-4) becomes

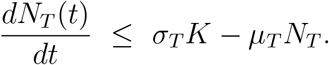

Thus,

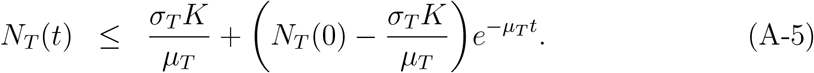

Equations (A-2) and (A-5) implies that *N*_*M*_ *T* (*t*) and *N*_*T*_ (*t*) are bounded and all solutions starting in the region Ω remain in Ω. Thus, the region is positively-invariant and hence, the region Ω attracts all solutions in 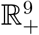.□

## B Stability of disease-free equilibrium (DFE) and the reproduction number *ℛ*_0_ of the Lyme disease model (5)

In this section, the conditions for the stability of the equilibria of the model (5) are stated. The Lyme disease model (5) has a disease free equilibrium (DFE). The DFE is obtained by setting the right-hand sides of the equations in the model (5) to zero, which is given by

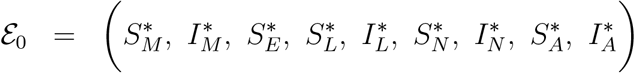

where

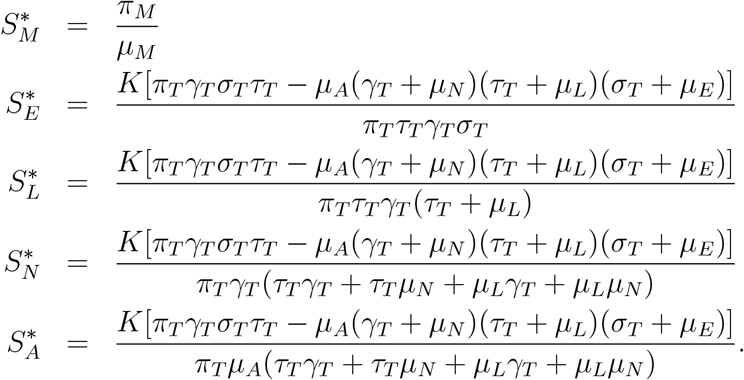

The stability of *ε*_0_ can be established by calculating the reproduction number *ℛ*_0_ using the next generation operator method on system (5). Taking *I*_*L*_, *I*_*N*_, *I*_*A*_, and *I*_*M*_ as the infected compartments and then using the notation in [22], the Jacobian *F* and *V* matrices for new infectious terms and the remaining transfer terms, respectively, are defined as:

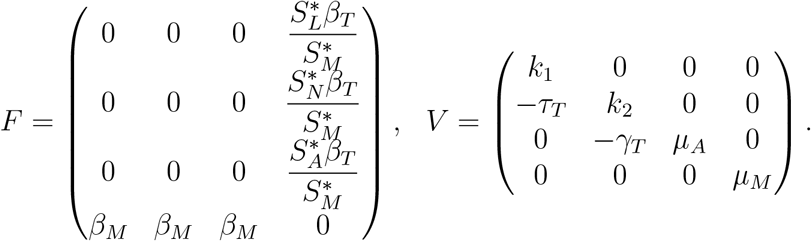

where *k*_1_ = *τ*_*T*_ + *μ*_*L*_, *k*_2_ = *γ*_*T*_ + *μ*_*N*_

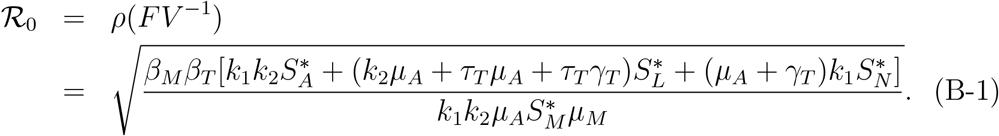

where, *ρ* is the spectral radius.

We made a simplifing assumption that *μ*_*L*_ = *μ*_*N*_ = *μ*_*A*_ = *μ*_*T*_. Then, the reproduction number in (B-1) becomes

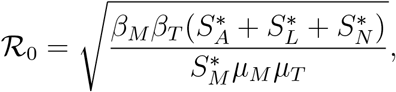

The expression *ℛ*_0_ is the number of secondary infections in completely susceptible population due to infections from one introduced tick or mouse with Lyme disease. Further, using Theorem 2 in [22], the following result is established.

### Lemma 3.

*The disease-free equilibrium (DFE) of the Lyme disease model* (5) *is locally asymptotically stable (LAS) if* ℛ_0_ < 1 *and unstable if* ℛ_0_ > 1.

## C Existence and Stability of Disease Free Periodic Solution

To determine the existence of the disease-free periodic solution, we first sum up the last seven equations of model (8), and let *μ*_*L*_ = *μ*_*N*_ = *μ*_*A*_ = *μ*_*T*_, and *ν*_*L*_ = *ν*_*N*_ = *ν*_*A*_ = *ν*_*T*_. Now, using the fact that *K* is the carrying capacity, it follows that *S*_*E*_ *≤ K*, hence, for *t ≥* 0 system (8) simplifies to

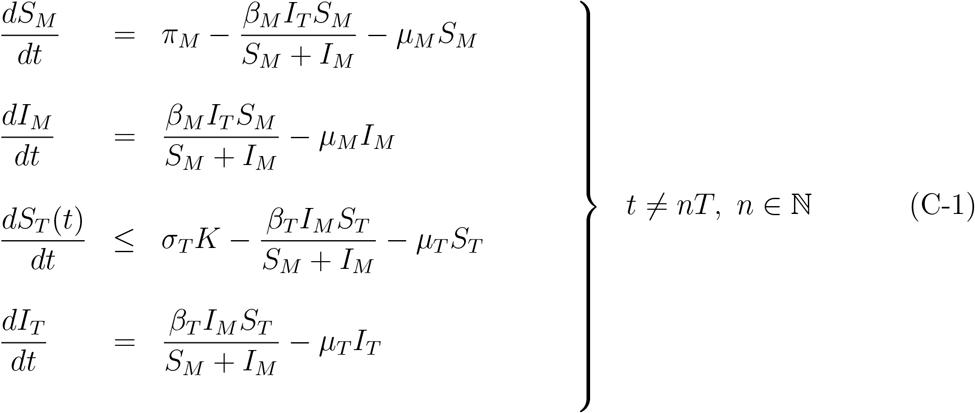

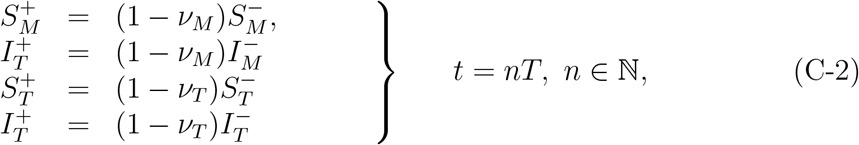

where *S*_*T*_ = *S*_*L*_ + *S*_*N*_ + *S*_*A*_ and *I*_*T*_ = *I*_*L*_ + *I*_*N*_ + *I*_*A*_.

At disease free equilibrium, *I*_*M*_ (*t*) = *I*_*T*_ (*t*) = 0, then (C-1) and (C-2) becomes

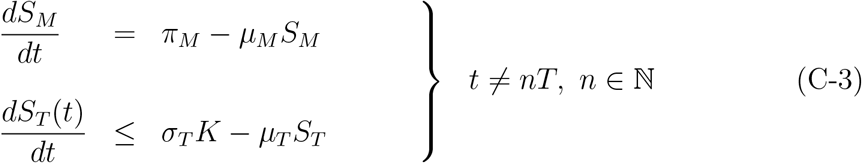

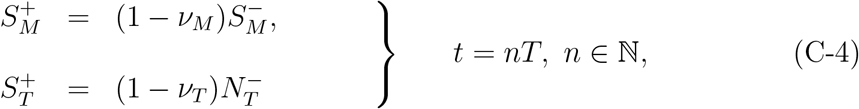

In the time interval *nT ≤ t≤* (*n* + 1)*T*, the first equation of system (C-3) has the solution

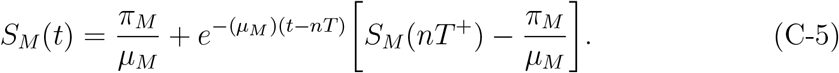

Let 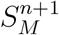 be the size of susceptible population after the (*n* + 1)-th pulse, i.e.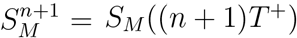. From (C-4) we have

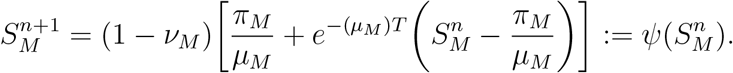

The map *ψ* has a unique positive fixed point

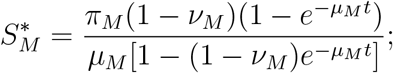

If *t ≠ nT*

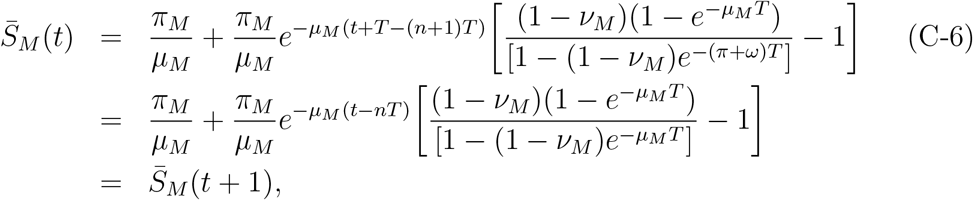

and in case *t* = *nT*, 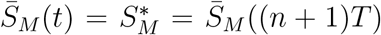, so (C-6) is periodic with period *T*. Thus, the solution of the first equation of (C-3) is a solution not only in the time interval [0, *T*), but also for all *t≥* 0. Hence, the solution of (C-6) in the time interval [0, *T*) is

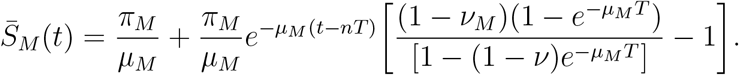

Following the above argument, the solution of the second equation of (C-3) for all *t ≥* 0 is given as

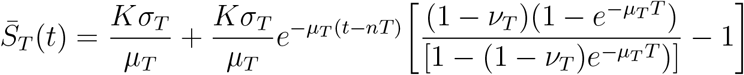

Further, similar to Gao et al. (2006, Lemma 2.2) [7], it can be shown that 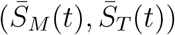 is globally asymptotically stable by using stroboscopic map. Hence, we summarize the results below as

### Lemma 4.

*The model* (C-3)(C-4) *has a unique disease-free periodic solution given as*

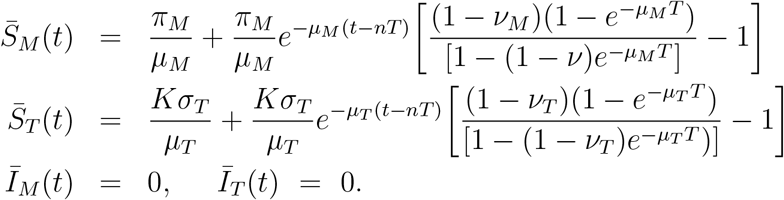

From Lemma 4, system (C-3)-(C-4), admits the disease-free periodic solution (DFPS) 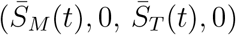 on every impulsive interval [*nT*, (*n* + 1)*T*]. To determine the stability of DFPS of system (C-3)-(C-4), we follow the approach in [24]. and define the following matrices

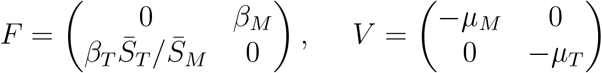

Let *A* be a *n* × *n* matrix, Φ_*A*(·)_(*t*) be the fundamental solution matrix of the linear ordinary differential system *x*′ = *Ax*, and *ρ*(Φ_*A*(·)_(*w*)) be the spectral radius of Φ_*A*(·)_(*w*). Let 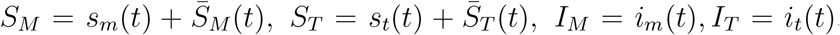. Then, system (C-3) can be written as

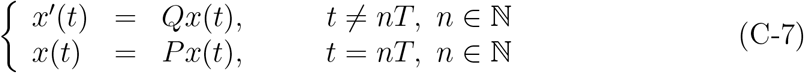

where

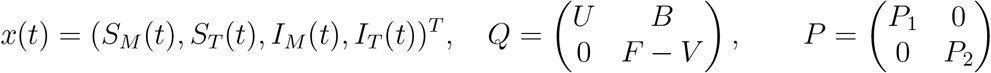

with

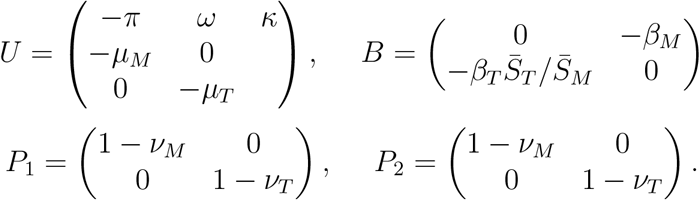

Let Φ_*Q*_(*t*) = (*Q*_*ij*_)_1*≤i,j≤*2_ be the fundamental matrix of *x*′(*t*) = *Qx*(*t*). Then Φ′_*Q*_(*t*) = *Q*Φ_*Q*_(*t*) with the initial value 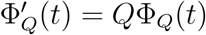, the identity matrix. Solving the equation gives

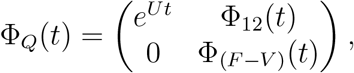

then we have

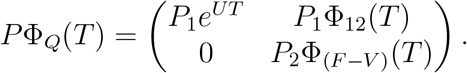

Therefore, the stability DFPS is dependent on eigenvalues of the matrices *P*_1_*e*^*UT*^ and *P*_2_Φ _(*F* −*V*)_ (*T*). The eigenvalues of *P*_1_*e*^*UT*^ are 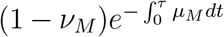, and 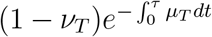, and we can see that the eigenvalues of *P*_1_*e*^*UT*^ are less that one. Furthermore, if spectral radius *ρ*(*P*_2_Φ_(*F* −*V*)_(*T*)) < 1, then DFPS is stable. This, leads to the following theorem.

### Theorem 1.

*If ρ*(*P*_2_Φ_(*F* −*V*)_(*T*)) < 1 *holds true, then the disease-free periodic solution* 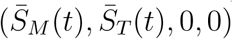 *system* (C-1)*-*(C-2) *is locally asymptotically stable*.

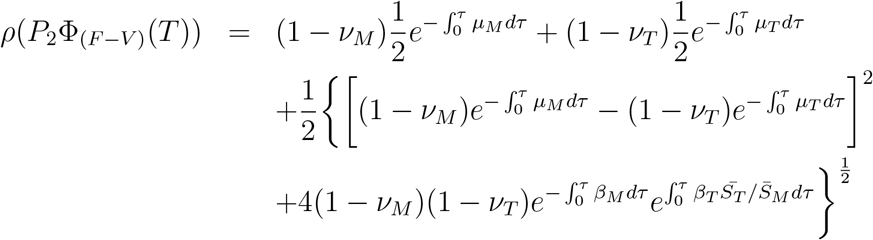

We denote the spectral radius as *ℛ*_*p*_ = *ρ*(*P*_2_Φ_(*F* −*V*)_(*T*)). Note, *ℛ*_*p*_ does not produce the number of individuals infected by a single infected carrier or infectious individual. Namely, it does not produce the average number of secondary infections [24]. However, it works as a threshold such that the disease persists as *ℛ*_*p*_ > 1 [24].

